# Microbial genes outperform species and SNVs as diagnostic markers for Crohn’s disease on multicohort fecal metagenomes empowered by artificial intelligence

**DOI:** 10.1101/2023.02.09.23285672

**Authors:** Sheng Gao, Xiang Gao, Ruixin Zhu, Dingfeng Wu, Zhongsheng Feng, Na Jiao, Ruicong Sun, Wenxing Gao, Qing He, Zhanju Liu, Lixin Zhu

## Abstract

**Background:** Dysbiosis of gut microbial community is associated with the pathogenesis of CD and may serve as a promising non-invasive diagnostic tool. We aimed to compare the performances of the microbial markers of different biological levels by conducting a multidimensional analysis on the microbial metagenomes of CD.

**Methods:** We collected fecal metagenomic datasets generated from eight cohorts that altogether include 870 CD patients and 548 healthy controls. The microbial alterations in CD patients were assessed at multidimensional levels including species-, gene- and SNV- level, and then diagnostic models were constructed using artificial intelligence algorithm.

**Results:** A total of 227 species, 1047 microbial genes and 21877 microbial SNVs were identified that differed between CD and controls. The species-, gene- and SNV- models achieved an average AUC of 0.97, 0.95 and 0.77, respectively. Notably, the gene model exhibited superior diagnostic capability, achieving average AUCs of 0.89 and 0.91 in internal and external validations, respectively. Moreover, the gene model was specific for CD against other microbiome-related diseases. Further, we found that phosphotransferase system (PTS) contributed substantially to the diagnostic capability of the gene model. The outstanding performance of PTS was mainly explained by genes *celB* and *manY*, which demonstrated high predictabilities for CD with the metagenomic datasets and was validated in an independent cohort by qRT-PCR analysis.

**Conclusions:** Our global metagenomic analysis unravels the multidimensional alterations of the microbial communities in CD, and identifies microbial genes as robust diagnostic biomarkers across geographically and culturally distinct cohorts.

## Introduction

Crohn ‘s disease (CD), one of the two main forms of inflammatory bowel disease (IBD), is characterized by skip lesions and transmural inflammation of the gastrointestinal tract. The incidence of CD has risen globally in past two decades, causing substantial economic burdens for patients and societies [1, 2]. Currently diagnosis of CD is mainly based on the combined evaluation of endoscopic, radiographic and pathological findings [3, 4]. However, the diagnostic power of endoscopy is often limited by patient compliance, bowel preparation quality and other uncontrollable factors [5]. Therefore, a sensitive, specific and convenient non-invasive diagnostic tool for CD is urgently needed.

Serologic and fecal biomarkers, such as C-reactive protein and fecal calprotectin, have been used as indicators to evaluate inflammatory activity in IBD [6, 7]. However, the accuracy and specificity of these biomarkers are not satisfactory. Recently, the diagnostic potential of the microbial signatures has emerged as potential diagnostic markers for IBD [8-12]. For instance, Pascal et al. constructed a diagnostic model using microbial species abundance and achieved a sensitivity of 81.8% for CD [11]. Similarly, Franzosa et al. reported a model that achieved an area under the ROC curve (AUC) of 0.92 [12]. Along this line, future effort is needed to conduct similar analysis incorporating multiple cohorts of distinct cultural and geographical background to identify markers of universal value.

Notably, species abundance may not be an accurate representative of the microbial functions as reflected by the fact that the nomenclatures of many gut microbial species are currently and constantly being adjusted. In this regard, the diagnostic value of microbial genes and their polymorphisms has become popular subjects of investigation [13-16] (**Fig. 1B**). For example, microbial functional genes outperformed microbial species in distinguishing CRC from controls [14]. Similarly, a recent study demonstrated high accuracy of microbial SNVs for diagnosing CD [17]. Currently, an integrated investigation on multidimensional signatures of CD at species-, gene- and SNV-levels is missing and seems to be warranted in the clinic.

**Fig. 1.**
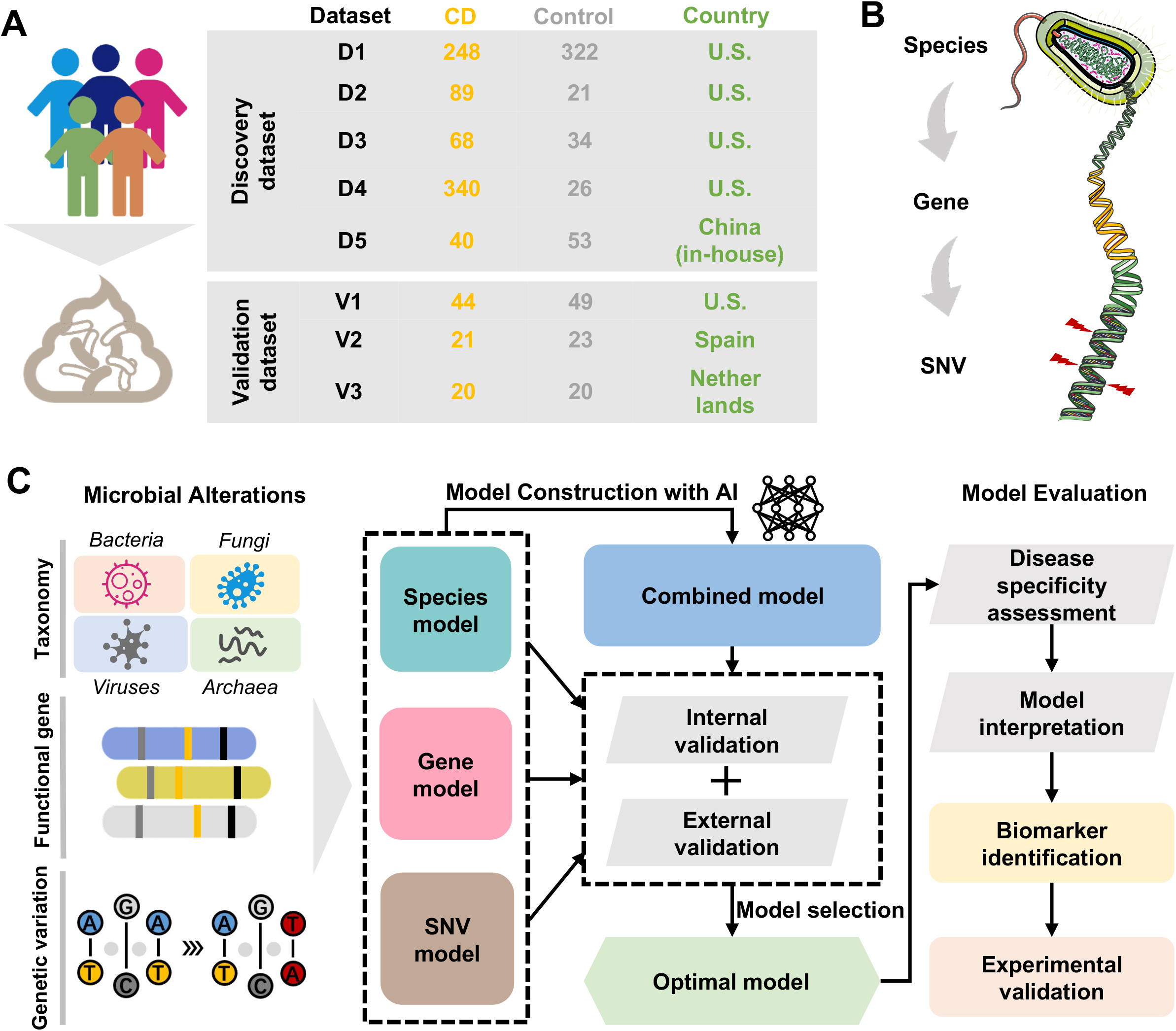
Overview of the fecal samples included in this study and the analysis protocol. **A** We collected a total of 1418 samples from 8 cohorts with fecal shotgun metagenomic data. The discovery dataset included D1, D2, D3, D4 and D5. The validation dataset included V1, V2 and V3. **B** Three levels of analysis were conducted in this study: species-, gene- and microbial SNV-levels. **C** The overall workflow of the study: Firstly, the microbial alterations were identified to retrieve the differential multidimensional signatures of gut microbiome. Subsequently, diagnostic models were constructed and the optimal model was selected according to the performances of the models in internal and external validations. Finally, disease specificity of model was evaluated and model interpretation was conducted for final determination of the microbial biomarker, and then biomarkers were validated by qRT-PCR analysis.

In this study, with large numbers of whole metagenome sequencing (WMS) samples from multiple cohorts, we constructed diagnostic models for CD and systematically assessed the predictabilities of multidimensional signatures. Candidate biomarkers for CD diagnosis were identified and further validated by qRT-PCR with an independent cohort. Collectively, these results uncover the multidimensional alterations of the microbial communities in CD patients and provide unbiased and robust biomarkers for CD diagnosis.

## Methods

### Study inclusion and data acquisition

For discovery dataset, we used PubMed to search for studies that published fecal shotgun metagenomic data of CD patients and controls. Raw FASTQ files of 1241 fecal samples from four studies were downloaded from the European Nucleotide Archive (ENA) including datasets ‘D1-D4 ‘ (**Fig. 1A)**.

For validation dataset, the raw data of 177 samples from three studies were collected from the ENA including datasets ‘V1-V3 ‘ (**Fig. 1A)**. The clinical characteristics of patients were shown in **Table S1**.

To evaluate whether the prediction model is specific for CD rather than non-CD diseases, we further collected five cohorts of non-CD diseases including ulcerative colitis (UC), colorectal cancer (CRC), type-2 diabetes (T2D), liver cirrhosis (LC) and Parkinson ‘s disease (PD).

### Patient recruitment and sample collection of Chinese cohorts

The Chinese cohort D5 consisted of 40 CD and 53 control samples (**Table S2**). The CD patients and controls were enrolled at the Sixth Affiliated Hospital of the Sun Yat-sen University, Guangdong province, China.

For qRT-PCR validation, we enrolled CD patients and controls at the Shanghai Tenth People ‘s Hospital. Patients with diagnosis of CD were included in the study. Potential participants were excluded if they were pregnant, were diagnosed with indeterminate colitis, had an acute gastrointestinal infection, or had antibiotic therapy within 3 months. In total, we collected 73 fecal samples (N = 37 for CD and N = 36 for control, **Table S3**) that were then stored at −80 °C before DNA extraction. The study was approved by the Institutional Review Board at the Shanghai Tenth People ‘s Hospital, Tongji University, Shanghai (No. 20KT863), and each participant provided informed consent.

### Quality control of WMS sequencing data

For preprocessing of the WMS sequencing data, quality control was performed using KneadData V0.6.0. Subsequently, reads with length lower than 50bp, or with low quality bases were filtered out by Trimmomatic software (V0.32). Furthermore, reads that mapped to the mammalian genome, bacterial plasmids, UNiVec sequences, and chimeric sequences were removed.

### Annotation and abundance estimation of microbial taxa, genes and SNVs

For multi-kingdom species level analysis, The customized reference database was constructed with 18756 bacterial, 359 archaeal, 9346 viral reference genomes from the NCBI Refseq database (accessed on January 2020), and 1094 fungal reference genomes from the NCBI Refseq database, FungiDB (http://fungidb.org) and Ensemble (http://fungi.ensembl.org) (all accessed on January 2020). Quality-filtered reads were aligned and quantified by Kraken2 [18] and Bracken, respectively.

For microbial gene level analysis, we assembled the quality-filtered metagenomes into contigs with Megahit (v1.2.9) [19] using ‘meta-sensitive ‘ parameters. Contigs shorter than 500-bp were excluded for further analysis. Prodigal (v2.6.3) software [20] was used to predict genes at the metagenome mode (-p meta). A non-redundant microbial gene reference was constructed with CD-HIT [21] using a sequence identity cut-off of 0.95, and a minimum coverage cut-off of 0.9 for the shorter sequences. The reference was annotated with EggNOG mapper (v2.0.1) based on EggNOG orthology data. Subsequently, CoverM (V4.0) was used to estimate gene abundances by mapping reads to the non-redundant reference and to calculate the coverage of genes in the original contigs. The abundance of KEGG orthologous (KOs) groups were calculated by summing the expression of genes annotated to the same KOs.

For SNV level analysis, MIDAS was used to perform microbial SNV annotation [22]. A customized reference genome database was constructed to include 7 species with sufficient coverage (>3X) in at least 20% of all samples. Then, the WMS reads were mapped to the database for SNV calling. Subsequently, the SNV profiles of all samples were merged, with only bi-allelic positions chosen. Other parameters were identical with those of the preset option ‘—core_snps ‘ (merge_midas.py snps –core_snps).

### Diagnostic model construction and evaluation

#### Model construction

Artificial intelligence (AI) algorithm called feedforward neural network (FNN) was employed to construct the diagnostic model. In detail, the hidden layers were activated by rectified linear unit (ReLU) activation function and the output layer was activated by sigmoid function. Subsequently, we performed stratified ten-fold cross-validation to avoid overfitting issue and model estimation using Scikit-learn 1.1.0. Finally, we trained the diagnostic model with well-optimized hyperparameter combinations with TensorFlow 2.8.0. The feature importance was evaluated with SHapley Additive exPlanations (SHAP) [23] to explain the output of machine learning model.

#### Model interpretation

To better interpret the compositions and corresponding contributions of features in model, we grouped KO genes by gene sets based on the priori knowledge of KEGG database. Subsequently, we randomly shuffled the abundance values of KO genes of a gene set in validation dataset, and performed predictions using the constructed diagnostic model. The decrease of AUC was considered as the importance of gene set to the diagnostic model. The above procedure was repeated for 50 times.

#### Evaluation of the model ‘s robustness and generalization

To test the robustness and generalization of selected optimal model among distinct cohorts, we performed cohort-to-cohort transfer and leave-one-cohort-out (LOCO) validation as described in our previous studies [24, 25]. For cohort-to-cohort transfer, diagnostic models were trained on one single cohort and validated on each of the remaining cohorts. For LOCO validation, one single cohort was set as the validation dataset while all other cohorts were pooled together as the discovery dataset.

### Disease specificity assessment of prediction model

Using non-CD diseases samples of UC, CRC, T2D, LC and PD, we evaluated the disease specificity of the predictive model for CD, following the method described by Thomas et al [26]. In detail, we randomly selected 10 control samples and 10 case samples from non-CD external data and added them into the control group in the validation dataset. If the model is specific for CD, the model would not perform worse with the addition of a case relative to the addition of the controls, because the model does not cover the characteristics of non-CD diseases. We repeated the procedure for 50 times.

### Validation of microbial genes by qRT-PCR

The gDNA was extracted with the TIANamp Stool DNA Kit (Cat# 4992205, TIANGEN) according to the manufacturer ‘s instructions. The primers used for validation are listed in **Table S4**. To perform the qRT-PCR analysis, the reaction mixture contained the primer pair with concentrations diluted to 0.2 μM and 10 ng gDNA in a 10 μl final volume with the SYBR Green qPCR Mix (Thermo Fisher Scientific). The cycling program was set as indicated: pre-denaturation at 95 °C for 10 min; denaturation at 95 °C for 15 s and annealing at 60 °C for 60 s for 40 cycles, followed by melting curve analysis. The qRT-PCR results were quantitated by calculating −ΔΔCt values between candidate genes and the 16S gene. The significance of the comparison between CD and control samples was tested by a two-sided Wilcoxon rank-sum test (*P* < 0.05).

### Statistical analysis

#### Alpha and beta diversity analysis

Alpha diversity of taxonomic profiles including Shannon, ACE, Simpson and Chao1 index were calculated based on Bray-Curtis distance using R (V4.0.5) “vegan” (V2.5.7) package. Beta diversity between groups were calculated by permutational multivariate analysis of variance (PERMANOVA) called adonis test, and significance was evaluated with 999 permutations.

#### Co-abundance analysis

Firstly, we generated species abundance profiles of CD and controls, respectively. Then we employed SparCC [27] to perform co-abundance analysis of differential multi-kingdom species. Correlations between differential multi-kingdom species were determined with 50 iterations. Then SparCC resampled the original dataset through bootstrap method to obtain random datasets. Later, pseudo-p-values are calculated from these random data sets to assess the significance of the initial observation scores. The statistical significance was calculated with 999 permutations. The network was visualized with Gephi (V0.9.5).

#### Multidimensional signatures association analysis

To further explore the potential associations between multi-dimensional signatures, Hierarchical All-against-All association testing (HALLA, V 0.8.20) [28] was performed. We generated species-, gene-, and SNV-profiles of CD patients and controls, respectively. Subsequently, the associations between the species-, gene-, SNV-signatures were calculated in pairs by HALLA. After that, we merged the output correlation matrices. Correlations with |cor| > 0.4 and *P*-values < 0.05 were used to constructed the network and visualized with Gephi (V0.9.5).

## Results

### Characterization of multicohort WMS data and study design

In this study, we collected eight fecal shotgun metagenomics datasets from published studies to characterize the gut microbiome in CD patients compared to healthy controls (**Fig. 1A**). Patients treated with antibiotics were excluded. In total, we included 785 samples from CD patients and 456 healthy control samples across geographically distinct regions from U.S. and China as the discovery dataset. In addition, 85 CD samples and 92 controls from three independent cohorts from U.S., Spain and Netherlands were included as the validation dataset. The overall protocol for this study (**Fig. 1C**) was based on the workflow of a previous study [24] with modifications.

### Multidimensional alterations in gut microbial profiles in CD patients

At species level, we found that alpha and beta diversities were significantly differed between CD patients and controls (**Fig. 2A-B**). A total of 80 bacterial species were identified with significantly different abundances between CD and control, such as *Escherichia coli, Flavonifractor plautii, Klebsiella pneumoniae* and *Bacteroides intestinalis*. (**Fig. 2C; Table S5**). Besides, 147 non-bacterial species including 70 fungus, 42 viruses and 35 archaea exhibited differential abundances between CD and controls, such as *Aspergillus rambellii, Capronia epimyces, Bacteroides phage B124-14, Klebsiella virus KpV80* and *DPANN group archaeon LC1Nh* (**Fig. S1 and Table S5**). Further, we investigated the differences in microbial interactions between CD and controls by performing co-abundance analysis via SparCC. Interestingly, interactions among intra-kingdom species were more frequently observed in the network of CD, compared to the network of controls (**Fig. S2**), indicating large scale alterations in the structure and function of the gut microbiome in CD.

**Fig. 2.**
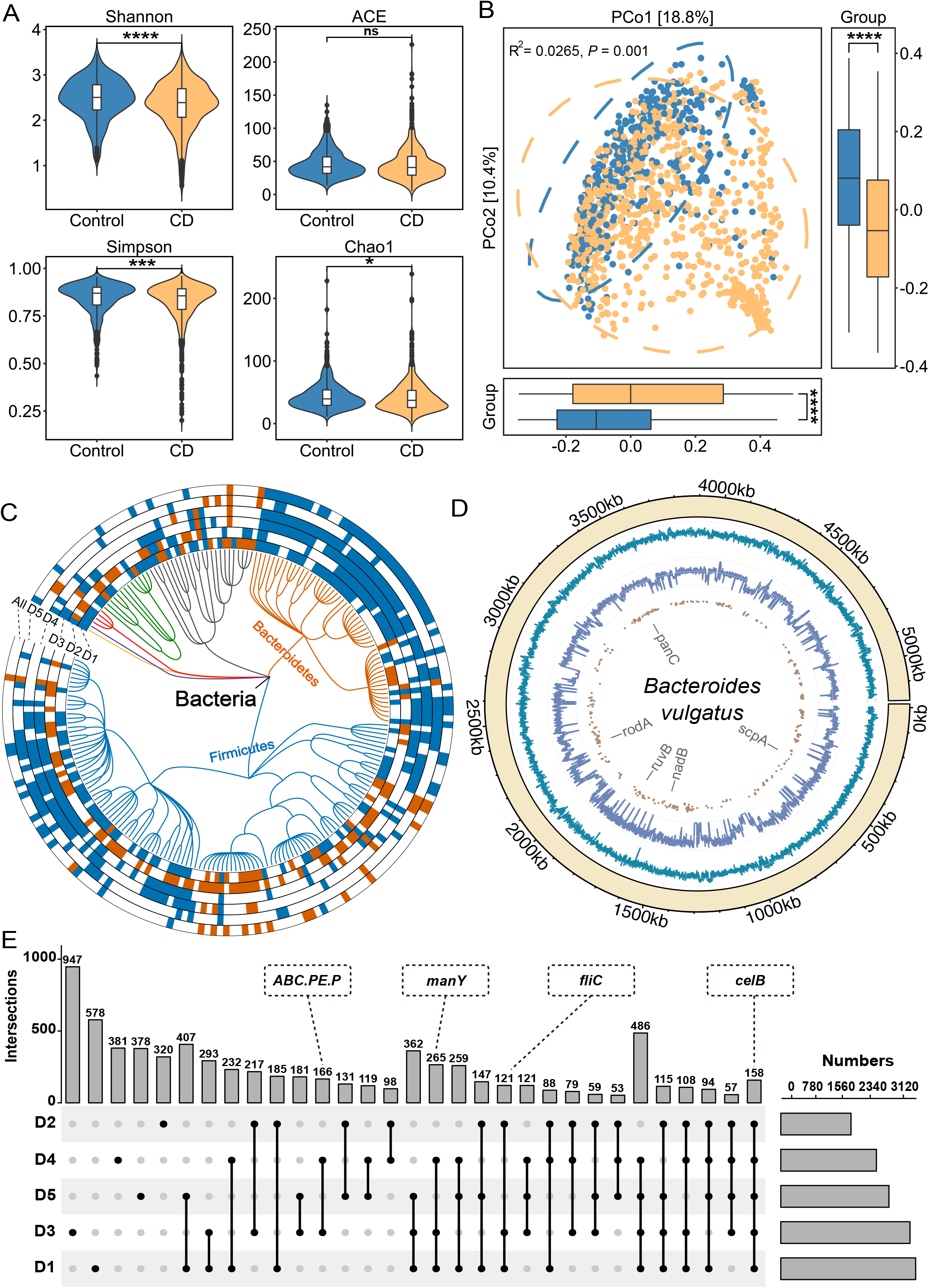
Multidimensional alterations in the gut microbiome of CD patients at species-, gene- and SNV-levels. **A** Alpha diversity measured by Shannon, ACE, Simpson and Chao1 index of patients with CD (orange, n = 785) and control individuals (blue, n = 456); \**P* < 0.05, \*\**P* < 0.01, \*\*\**P* < 0.001 and \*\*\*\**P* < 0.0001. **B** Principal coordinate analysis (PCoA) of samples from all five cohorts based on Bray–Curtis distance, which shows that microbial compositions were different between groups (R^2^ = 0.0265, *P* = 0.001). P values of beta diversity based on Bray–Curtis distance were calculated with PERMANOVA by 999 permutations (two-sided test). **C** Phylogenetic tree showing the differential bacteria species, grouped by the phyla. The differential species in each dataset are shown in each circle ‘D1-D5 ‘ (*P* < 0.05, two-sided test); the meta-analysis results in integrated dataset were marked by ‘All ‘. Increased and decreased abundances are indicated by red and blue, respectively. **D** The chord diagram shows the distributions of annotated SNVs in *Bacteroides vulgatus* genome. The outer circle represents the genome of *B. vulgatus*; the inner circles represent the GC-content (cyan indigo lines), sequencing depth (purple lines) and sites of differential SNVs (brown points) in the genome, respectively. **E** UpSet plot showing the number of differential KO genes identified via MaAsLin2 in each dataset and those shared by the datasets. The number above each column represents the intersection size of differential KO genes. The connected dots represent the common differential genes across connected cohorts. The set size on the right represents the number of differential genes in each cohort.

Next, we assessed the microbial alterations at KO gene level, and identified 497 genes with increased abundance and 1043 genes with decreased abundance in CD patients, such as the genes encoding peptide/nickel transport system permease protein (*ABC*.*PE*.*P*), mannose PTS system EIIC component (*manY*), flagellin (fliC) and cellobiose PTS system EIIC component (*celB*) (**Fig. 2E; Table S6**). For better understanding of these differential KO genes, we performed gene set enrichment analysis (GSEA). 59 enriched pathways, including 18 pathways with increased abundances and 41 with decreased abundances in CD patients, were identified (**Fig. S3A and Table S7**). Propanoate metabolism, quorum sensing, phosphotransferase system (PTS) and purine metabolism exhibited increased abundances in CD, while biosynthesis of secondary metabolites, pantothenate and CoA biosynthesis exhibited decreased abundances in CD.

For microbial SNV level analysis, a total of 7 commonly observed species that have sufficient coverage (> 3X) in at least 20% of the samples were annotated, with the number of SNVs ranging from 74 with *Bacteroides rodentium* to 99305 with *Bacteroides vulgatus* (**Fig. S3B and Fig. S4**). A total of 21877 differential SNVs were identified in the seven annotated species (**Fig. S3C**). For instance, *Bacteroides vulgatus*, belonging to the most commonly encountered *Bacteroides* species in the human colon, had 11134 significantly differential SNVs that located on genes such as *panC, rodA* and *ruvB*. (**Fig. 2D; Table S8**). These differential SNVs are potential candidates of risk factors mediating abnormal gene functions. Collectively, we systemically assessed the multidimensional microbial alterations in CD patients compared to controls, and identified differential signatures for diagnostic model construction.

### Diagnostic models for CD based on microbial multidimensional signatures

Based on all of the differential signatures at species-, gene- and microbial SNV-levels, we constructed models using deep learning algorithm. At species level, we firstly evaluated the capability of single-kingdom species for distinguishing CD from controls. The average AUCs of cross-validation based on fungal, viral, archaeal signatures were 0.89, 0.81 and 0.76, respectively. Compared to non-bacterial species, bacterial species demonstrated a better performance in disease prediction (average AUC=0.94) (**Fig. S5A-D**). Furthermore, we merged single-kingdom signatures together, and found that the species model based on multi-kingdom signatures had higher diagnostic accuracy with an average AUC of 0.97 (**Fig. 3A; Fig. S5E**). Interestingly, we noticed that several fungal species including *A. rambellii* and *A. ochraceoroseus* were top-ranking features of the model with high SHAP values, suggesting their close association with CD pathology (**Fig. S6A and Table S9**).

**Fig. 3.**
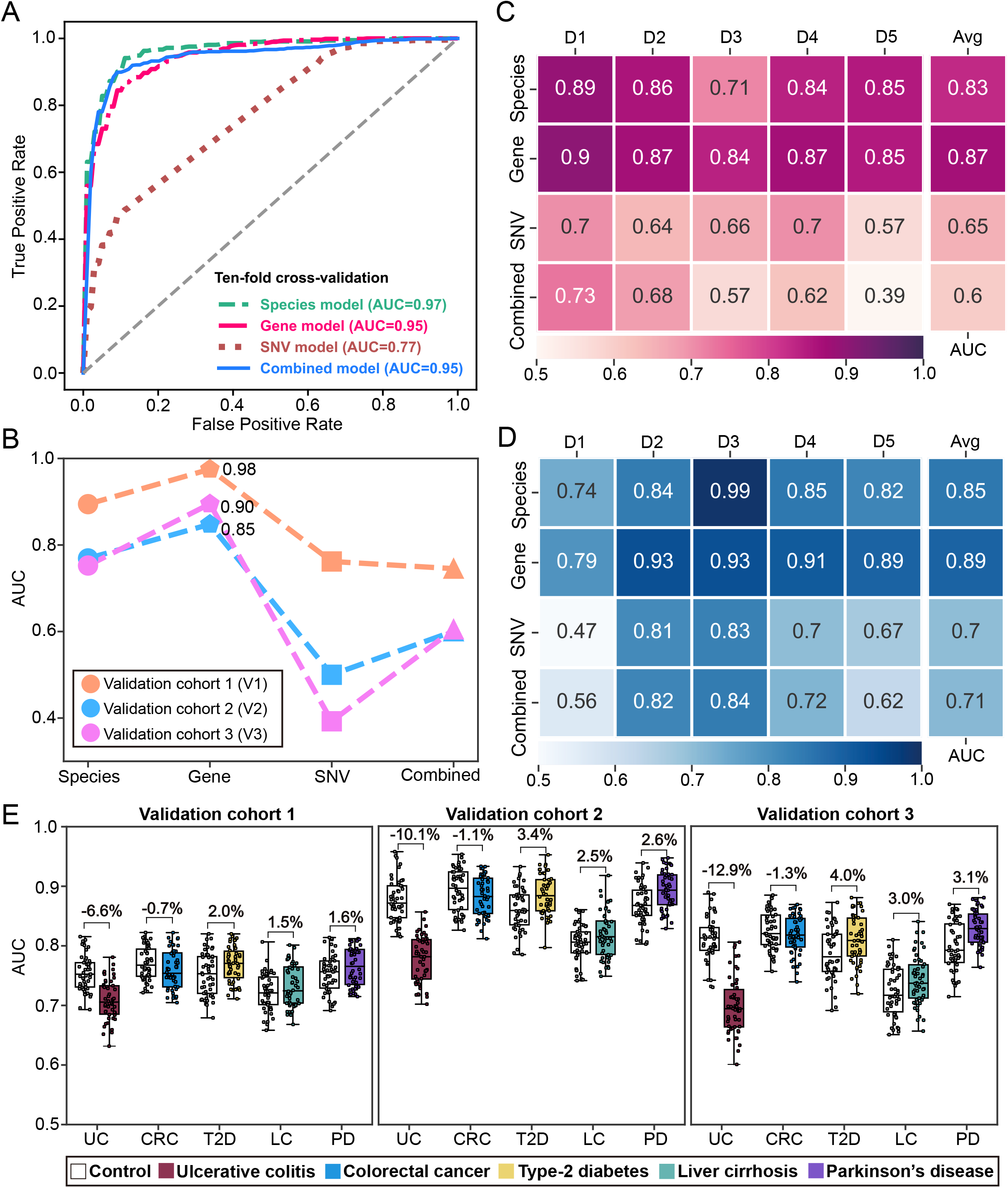
The performance of diagnostic models constructed with multidimensional signatures. **A** The ROC curves from ten-fold cross-validation of species-, gene-, SNV- and combined diagnostic models. **B** The AUCs of species-, gene-, SNV- and combined diagnostic models in external validation dataset. **C** The AUCs of each model in cohort-to-cohort validation. Each number represents the average AUC of validation with the cohort specified by its column tag as the training cohort, and all other cohorts as the validation cohorts. **D** The AUC of each model in LOCO validation. Each number represents the resulting AUC of validation with the cohort specified by its column tag as the validation cohort while the other cohorts combined as training cohort. **E** Prediction performances as AUC values on the validation cohorts when adding an external set of control and case samples from non-CD disease cohorts (ulcerative colitis (UC), colorectal cancer (CRC), type-2 diabetes (T2D), liver cirrhosis (LC) and Parkinson ‘s disease (PD)). Gray and colored bars are the AUCs after adding control and case samples from the non-CD disease cohorts, respectively.

Subsequently, we constructed a diagnostic model based on 1047 differential KO genes. The gene model achieved an average AUC of 0.95 in 10-fold cross-validation, slightly lower than that of the multi-kingdom species model (**Fig. 3A**). From feature importance evaluation, we found that CDP-abequose synthase (*rfbJ*), type VI secretion system protein ImpB (*impB*), nitrite reductase (NO-forming) (*nirK*), and *celB* were the most important KO genes with SHAP values ranged from 0.006 to 0.008 (**Table S10**). Notably, the KO gene *celB* was found to be significantly increased in CD patients of each dataset (**Fig. S6B**), suggesting an outstanding contribution of *celB* gene to the diagnostic power of the model.

Furthermore, we explored the diagnostic potentials of microbial SNVs. The SNV model achieved an average AUCs of 0.77 in cross-validation (**Fig. 3A**). The most important SNVs were mainly from *Bacteroides* species including *B. ovatus, vulgatus* and *uniformis* (**Fig. S7A and Table S11**). As the most widely colonized microbes in the gut [29], *Bacteroides* species contributes to the major diagnostic power of the SNV model in our results.

Finally, we constructed a model with the combination of species-, gene- and SNV-signatures **(Fig. S8D**). The combined model achieved an average AUC of 0.95. Interestingly, the performance of combined model was not significantly improved compared to species- and gene-models, and most of the top-ranking features were from KO genes (**Fig. 3A; Fig. S7B**). These results suggest that the gene signatures are the most powerful biomarkers for CD.

### Gene model achieves superior robustness and generalization

To assessed the robustness and generalization of species-, gene-, SNV- and combined models, we performed internal and external validations. With the internal validation cohorts, the gene model achieved the highest average AUCs of 0.87 and 0.89 in cohort-to-cohort transfer and LOCO validation, respectively (**Fig. 3C-D**), compared to other diagnostic models (**Fig. S9A-G**). In external validation, the gene model also exhibited the best performance with an average AUC of 0.91 in three independent cohorts (**Fig. 3B**). Taken together, the gene model demonstrated superior robustness compared to the species-, SNV-model and even the combined model.

### Gene model is highly specific for CD

To ascertain the discriminative power of the gene model, that is, the model is specific for CD but not other microbiome-related diseases, we chose five microbiome-related diseases including UC, CRC, PD, T2D and LC to evaluate the disease specificity of the gene model. Adding UC samples into three independent validation cohorts decreased the AUC by 6.6, 10.1 and 12.9%, respectively (**Fig. 3E**). These changes were not significant considering the baseline values of the altered AUCs when adding CD samples in the validation dataset (decreased AUCs by 10.7, 17.5 and 20.5%, respectively, **Fig. S9H**). With CRC cohorts, slight and insignificant changes of AUCs in validation (decreased by 0.7, 1.1 and 1.3%, respectively) were observed. Similarly, slight and insignificant changes of AUC were observed in validations with T2D (increased by 2.0, 3.4 and 4.0%, respectively), liver cirrhosis (increased by 1.5, 2.5 and 3.0%, respectively) and PD (increased by 1.6, 2.6 and 3.1%, respectively). Altogether, the slight changes in AUCs suggest limited effects of the samples with non-CD diseases on the CD model, indicating that our diagnostic model is specific for CD.

### Outstanding contributions of phosphotransferase system to the diagnostic capability of the gene model

To evaluate the respective contributions of each gene set and of key gene feature in the gene model, the KO gene features were grouped by gene set and the importance of each gene set was evaluated as described in Methods section. Relative to the baseline AUC of 0.91, the abundance disturbance of the gene sets quorum sensing, PTS and ABC transporters caused the greatest decrease of AUC in the predictive model by 1.09 to 1.70 percent (**Fig. 4A**). Further, we performed recursive feature elimination by gene sets and reconstructed diagnostic models. We found that the AUC of cross-validation did not decrease significantly until the glycerolipid metabolism gene set was eliminated, which confirmed the important contribution of quorum sensing, PTS, ABC transporters, fructose and mannose metabolism and glycerolipid metabolism to the diagnostic model (**Fig. S10A**). To further strengthen these results, we constructed a sub-model with genes of these five gene sets, which achieved an AUC of 0.89 in cross-validation (**Fig. S10B**). The sub-model displayed decent robustness in internal validations and achieved an average AUC of 0.81 in external validation (**Fig. S10C**). Notably, we found that *celB* was the most important feature in the sub-model (**Table S12**). These results suggest that the above identified gene sets are the key contributors to diagnostic capabilities of the gene model.

**Fig. 4.**
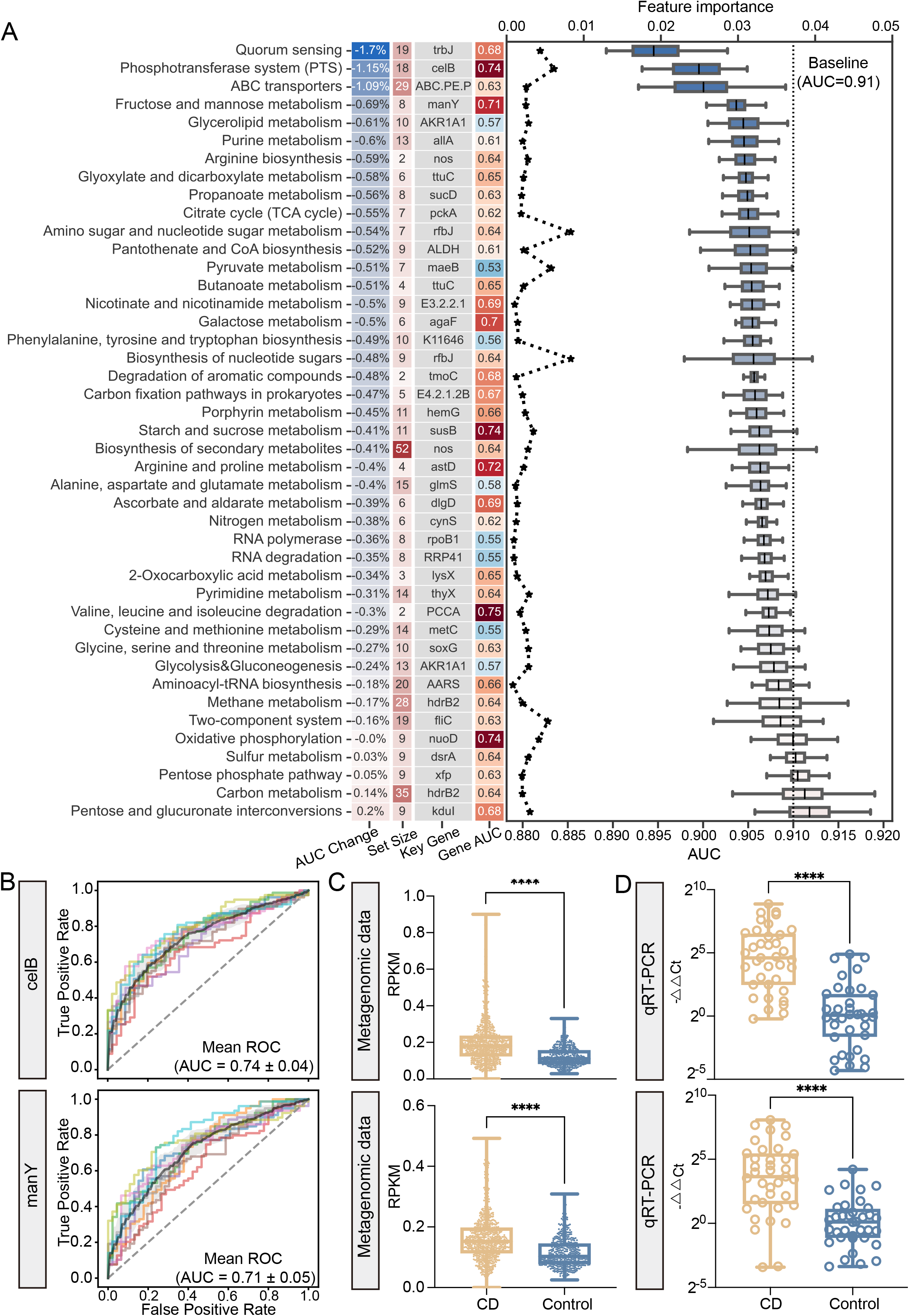
The model interpretation of the gene model. **A** The left column lists the average percent change of AUC after shuffling the abundance values of the genes in each gene set in validation dataset with the background color indicating the degrees of AUC change; the center left column lists the number of KO genes in each gene set with the background color indicating the set size; the center right column is the representative signature of each gene set; and the right column lists the cross-validation AUC of the representative microbial gene with the background color indicating an increased (red) or decreased (blue) AUC. The line plot shows the values of feature importance of the representative signatures (upper horizontal axis); the box plot shows the AUCs of each gene set in validation dataset with the dotted line representing the baseline AUC of 0.91 (lower horizontal axis). **B** The ROC curve shows diagnostic performance of microbial gene *celB* and *manY*, respectively. **C-D** The box plot shows the abundances of *celB* (upper) and *manY* (lower) in metagenomic data (C) and qRT-PCR data (D) (N=37, CD; N=36, control), respectively. Data are presented as mean ± standard deviation. \**P* < 0.05, \*\**P* < 0.01, \*\*\**P* < 0.001 and \*\*\*\**P* < 0.0001.

Next, we assessed the prediction power of representative KO genes of each gene set (**Table S13**). Notably, *celB* and *manY* displayed excellent diagnostic capabilities with AUCs of 0.74 and 0.71, respectively (**Fig. 4B**). Since *celB and manY* (also a member of fructose and mannose metabolism) are both members of PTS, the above results indicate that PTS gene set mediated the most significant functional alterations of gut microbiome in CD patients. Finally, we validated the abundances of *celB* and *manY* with an independent cohort of CD patients and controls by qRT-PCR. Consistent with the metagenomic data (**Fig. 4C**), both *celB* and *manY* were significantly more abundant in CD patients (**Fig. 4D**). Additionally, we validated the abundances of those genes that belong to important pathways and with high feature importance by qRT-PCR (**Fig. S11**). These results revealed the respective contributions of individual gene feature to the diagnostic capability of the gene model, and identified *celB* and *manY* as the individual biomarkers with the highest predictive power for diagnosing CD.

### Altered interactions within and between each level of microbial signatures in CD

For a global understanding of the interactions among all the microbial signatures in CD, we investigated the associations among all the microbial signatures via HALLA (**Fig. 5A-B**). In both CD and control networks, considerable associations were observed between KO genes and species, but few observed between SNVs and the other two levels (|correlation| > 0.4) (**Fig. 5B, E**). More associations were observed in the network of CD (206 associations) (**Fig. 5D)**, than in the network of controls (163 associations) (**Fig. 5G; Table S14-15**). Interestingly, there were more negative associations between the gene- and the species-signatures in the control network than that in the CD network. For example, D-nopaline dehydrogenase (*nos)*, type IV secretion system protein TrbJ (*trbJ*) genes were negatively associated with *R. hominis, R. bassiana*, and *C. aerofaciens*. Notably, we found that KO genes had stronger degree centrality than species in the CD network (**Fig. 5C**). Moreover, compared to the control network, these KO genes in CD tended to form isolated clusters, as exemplified by the independent module consisting *celB* and *manY* in the CD network (**Fig. 5A**).

**Fig. 5.**
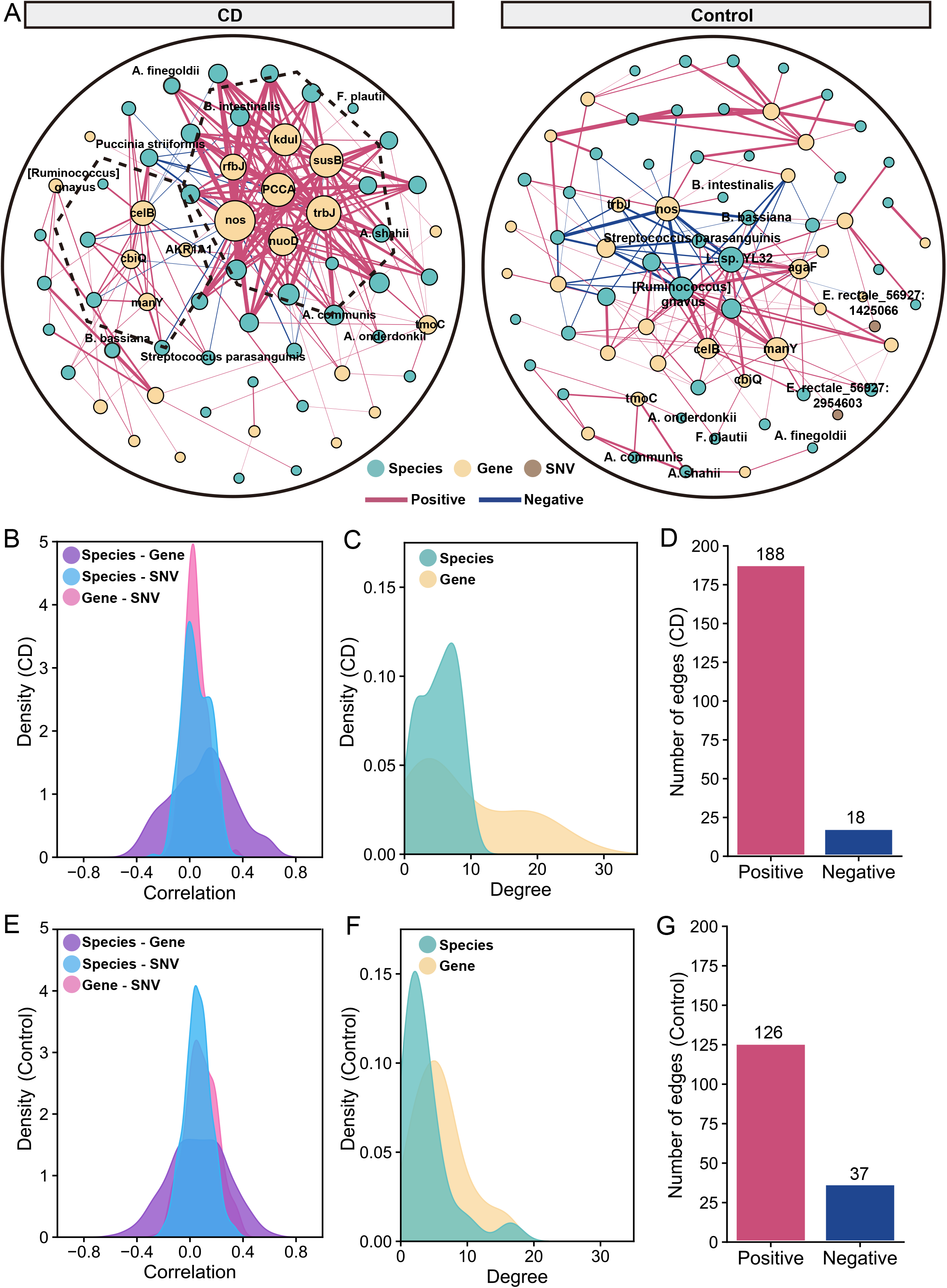
The cross-talk between multidimensional signatures. **A-B** Correlations among species-, gene- and SNV-signatures in the control (A) and the CD (B) networks. The colors of nodes indicate signatures of different levels: species (green), KO genes (yellow), and SNVs (brown). Red line indicates positive interaction; and blue line indicates negative interaction (|correlation|>0.4, FDR < 0.05). **C** Density distribution of correlations between different levels of signatures (FDR < 0.05) in the control network. **D** Density distribution of node degrees for different levels of signatures ((|correlation|>0.4, FDR < 0.05) in the control network. **E** Numbers of edges (|correlation|>0.4, FDR < 0.05) in the control network. **F** Density distribution of correlations between different levels of signatures (FDR < 0.05) in the CD network. **G** Density distribution of node degrees for different levels of signatures ((|correlation|>0.4, FDR < 0.05) in the CD network. **H** Number of edges (|correlation|>0.4, FDR < 0.05) in the CD network.

## Discussion

Here, for the first time, multidimensional microbial signatures of CD were systematically analyzed with multiple cohorts of distinct cultural and geographical backgrounds. In comparison of the diagnostic capabilities of the microbial signatures including differential species, genes and SNVs, the gene model achieved superior accuracy and robustness in distinguishing CD from controls, and the gene model was specific for CD against other microbiome-related diseases. Finally, the major contributing genes in the gene model were identified and validated.

The multidimensional alterations of the gut microbiome in CD patients contain massive information that could predict the disease state. Therefore, we employed deep learning method to fit the underlying characteristics of gut microbiome in CD. With the microbial species models, while bacterial species achieved the best performance among single-kingdom models, multi-kingdom model with both bacteria and non-bacterial species achieved better accuracy than the single-kingdom models, which is similar to our observations with the microbial models for colorectal cancer [14].

Comparing models of three different types, the gene model demonstrated the best generalization and robustness in model evaluations compared to the species-, SNV- and combined models. This is reasonable, considering that the homologous genes of different microorganisms may contribute to the same abnormalities in the gut microbiome in connection to specific pathological processes [30, 31].

In examining the contributions of individual gene set and gene to the diagnostic capabilities of the gene model, we found that genes that belong to PTS gene set had great impacts on the model accuracy in abundance disturbance analysis. The importance of the PTS gene set in the diagnosis model was also demonstrated in recursive feature elimination analysis and in cross-validation of the sub-model. In gut bacteria, the PTS is known as a system that catalyze sugar transport as well as sugar phosphorylation [32, 33]. In addition, the PTS regulates a wide variety of transport, metabolic processes, biofilm formation and virulence [34], thus it is considered as a comprehensive regulation and coordination system. We observed that the CD patients exhibited increased abundance in PTS, and that the KO genes in PTS were associated with the differential species in CD (**Fig. S12A**), such as *Streptococcus pneumoniae* and *A. ochraceoroseus* that are associated with gut diseases [35-37]. That is, PTS may participate in the pathogenesis of CD.

More importantly, the KO gene *celB* that encodes the enzyme IIC component (EIIC) of cellobiose PTS, exhibited the top-ranking predictability among all the gene markers and an increased abundance in CD patients. These observations support an outstanding potential for the microbial gene *celB* of PTS to be used as a biomarker for non-invasive CD diagnosis. Moreover, *celB* was associated with *K. pneumonia* (**Fig. S12B**), which is in line with the roles of *celB* component of PTS in biofilm formation and the virulence of *K. pneumoniae* [38], and the roles of *K. pneumoniae* in the initiation and perpetuation of the pathological damage of CD were also demonstrated [39]. We also observed a significant increase of *K. pneumoniae* in CD patients (**Fig. S12C and Table S5**). Therefore, it is reasonable to hypothesize that the interaction between *celB* and *K. pneumoniae* may contribute to the development of CD.

Another microbial gene *manY* was also identified as a biomarker for CD diagnosis. *manY* encodes the EIIC component of mannose PTS system (man-PTS) that is a part of the PTS regulatory network. The hairpin tips of IIC in man-PTS is coordinated with mannose and mediates the mannose transport [40]. Interestingly, previous studies found that man-PTS and cellobiose-PTS were upregulated in gut microbes by changing from a low-fat diet to a high-fat, high-sugar diet [40, 41], suggesting that the PTS of gut microbes is sensitive to the nutritional environment of mucosal surfaces. Thus, the up-regulations of *celB* and *manY* in CD likely indicate the up-regulation in the biological activities of cellobiose-PTS and man-PTS in association with CD pathology. That is, *manY* may also participate in the pathogenesis of CD. However, the cause of these alterations in CD is not clear and requires further investigation.

Our work takes advantages of excellent adaptability and learning ability of AI in large dataset and provides an effective non-invasive diagnostic tool with improved discrimination power for CD. However, the interpretability of the AI model is limited, which could be improved with a causal analysis in the future.

## Supporting information

Supplementary Figues

## Data Availability

All data produced in the present study are available upon reasonable request to the authors

## Conclusions

Our global metagenomic analysis unravels the multidimensional alterations of the microbial communities in CD and identifies microbial genes as robust diagnostic biomarkers across cohorts. These genes are functionally related to the pathogenesis of CD. Future study on these genes may lead to an effective non-invasive diagnostic tool for CD.

## Ethics approval and consent to participate

All participants provided written informed consent prior to data collection. The study was approved by the Institutional Review Board at the Shanghai Tenth People ‘s Hospital, Tongji University, Shanghai (No. 20KT863).

## Consent for publication

Not applicable.

## Availability of data and materials

The data that support the findings of this study are available from the corresponding author, upon reasonable request. The code and scripts are available on GitHub (https://github.com/tjcadd2020/Diagnosis-for-CD).

## Authors ‘ contributions

LZ, ZL, QH and RZ conceived and designed the project. SG and XG drafted the manuscript. RZ, DW, ZF, NJ, RS, WG, QH, ZL and LZ revised the manuscript. All authors read and approved the final manuscript.

## Acknowledgements

This work was supported by the National Natural Science Foundation of China (82170542 to RZ, 92251307 to RZ, 32200529 to DW, 82000536 to NJ), the National Key Research and Development Program of China (2021YFF0703700/2021YFF0703702 to RZ), and Guangdong Province “Pearl River Talent Plan” Innovation and Entrepreneurship Team Project (2019ZT08Y464 to LZ). The funders had no role in study design, data collection and analysis, decision to publish, or preparation of the manuscript.

## Abbreviations

ABC.PE.P: peptide/nickel transport system permease protein;
agaF: N-acetylgalactosamine PTS system EIIA component;
AKR1A1: alcohol dehydrogenase (NADP+);
ALDH: aldehyde dehydrogenase (NAD+);
allA: ureidoglycolate lyase;
AUC: area under the ROC curve;
CD: Crohn ‘s disease;
celB: cellobiose PTS system EIIC component;
CRC: colorectal cancer;
EIIC: enzyme IIC component;
ENA: European Nucleotide Archive;
fliC: flagellin;
FNN: Feedforward neural network;
GSEA: gene set enrichment analysis;
IBD: inflammatory bowel disease;
impB: type VI secretion system protein ImpB;
KO: KEGG Orthology;
LC: liver cirrhosis;
LOCO: leave-one-cohort-out;
maeB: malate dehydrogenase (oxaloacetate-decarboxylating) (NADP+);
manY: mannose PTS system EIIC component;
nirK: nitrite reductase (NO-forming);
pckA: phosphoenolpyruvate carboxykinase (GTP);
PD: Parkinson ‘s disease;
PTS: phosphotransferase system;
ReLU: rectified linear unit;
rfbJ: CDP-abequose synthase;
ROC: receiver operating characteristic;
SHAP: SHapley Additive exPlanations;
SNVs: single nucleotide variants;
sucD: succinyl-CoA synthetase alpha subunit;
T2D: type-2 diabetes;
tcPp: toxin coregulated pilus biosynthesis protein P;
tmoC: toluene monooxygenase system ferredoxin subunit;
trbJ: type IV secretion system protein TrbJ;
ttuC: artrate dehydrogenase/decarboxylase / D-malate dehydrogenase;
UC: ulcerative colitis;
WMS: whole metagenome sequencing

