## Supplementary Figues for "Microbial genes outperform species and SNVs as diagnostic markers for Crohn’s disease on multicohort fecal metagenomes empowered by artificial intelligence"

### Supplementary Materials

#### Supplementary Figures

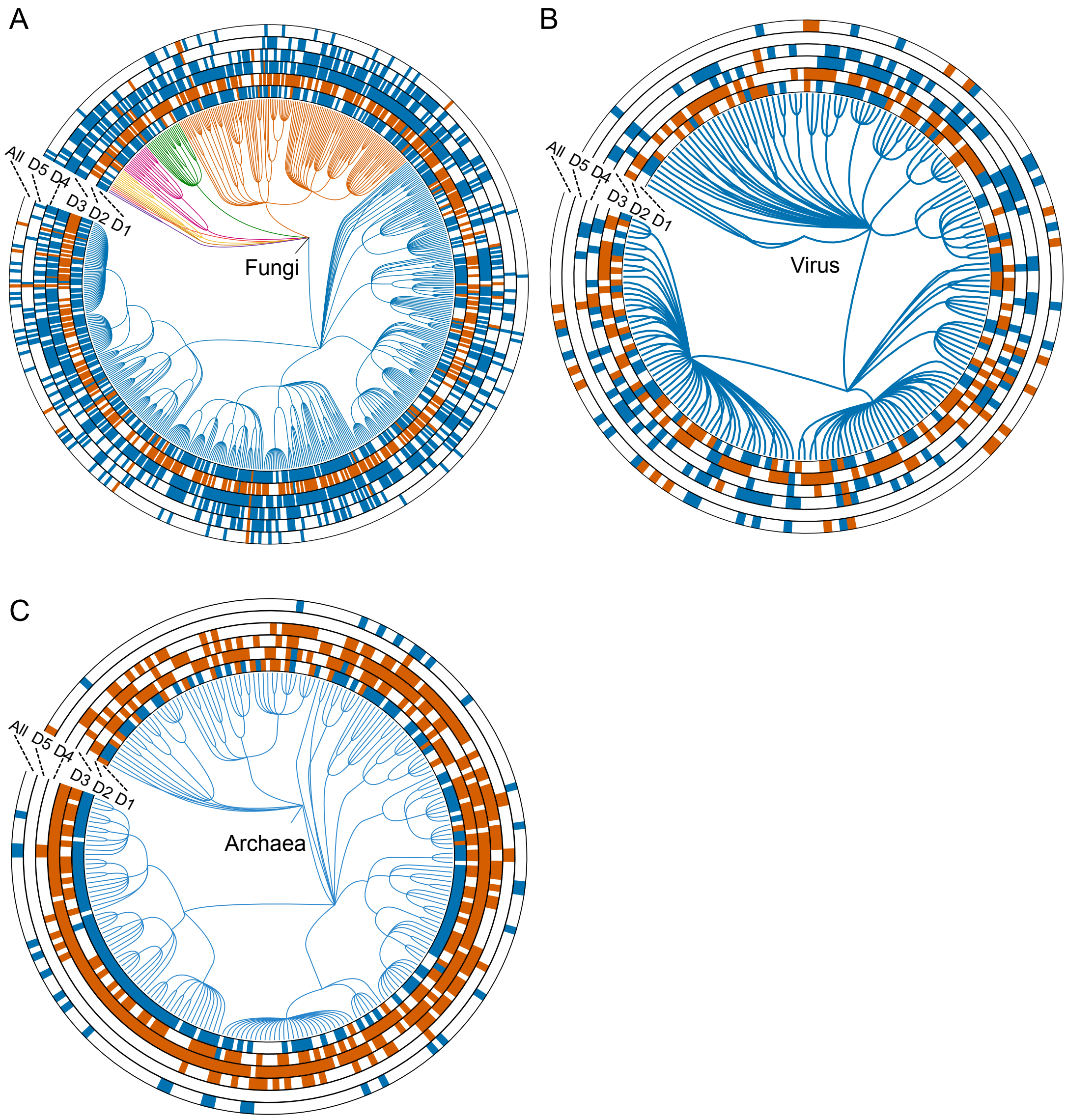


**Fig. S1. Differential taxa in non-bacterial kingdoms in CD patients. A-C** Phylogenetic tree showing the differential fungi (A), viruses (B) and archaea (C), respectively. The differential species in each dataset are shown in circles 'D1-D5' ($P < 0.05$, two-sided test); the meta-analysis results in integrated dataset were marked by 'All'. Increased and decreased abundances are indicated by red and blue, respectively.

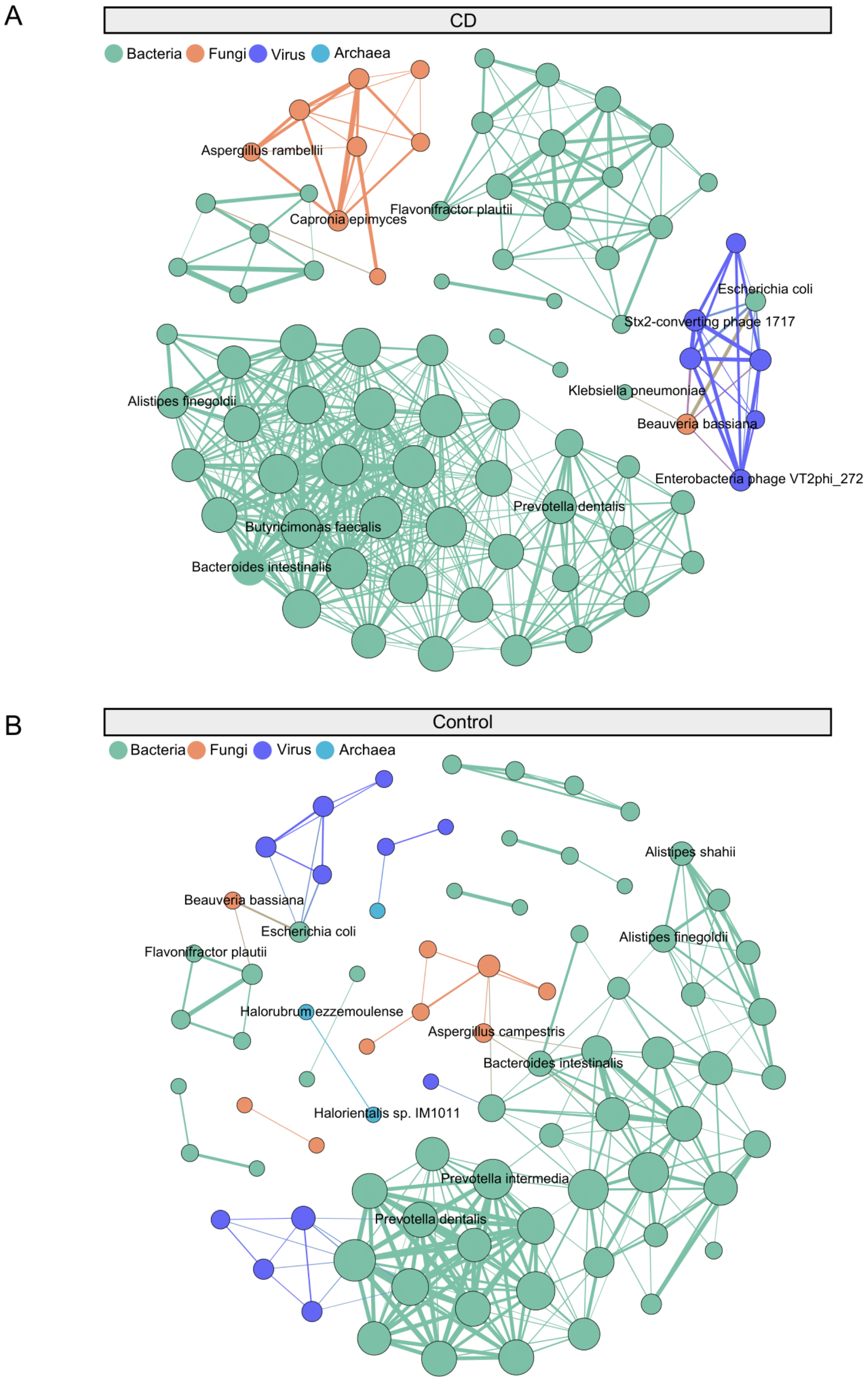


**Fig. S2. Correlations among multi-kingdoms microbes in CD patients and controls. A-B** Multi-kingdom microbial correlation networks of CD (A) and controls (B) by SparCC (|cor| > 0.6, FDR < 0.00001, two-sided tests of 1,000 permutations). Bacteria, fungus, viruses and archaea are colored by green, orange, purple and blue, respectively. Node size represents node degree in the network, and the line width represents the weight of correlations between nodes.

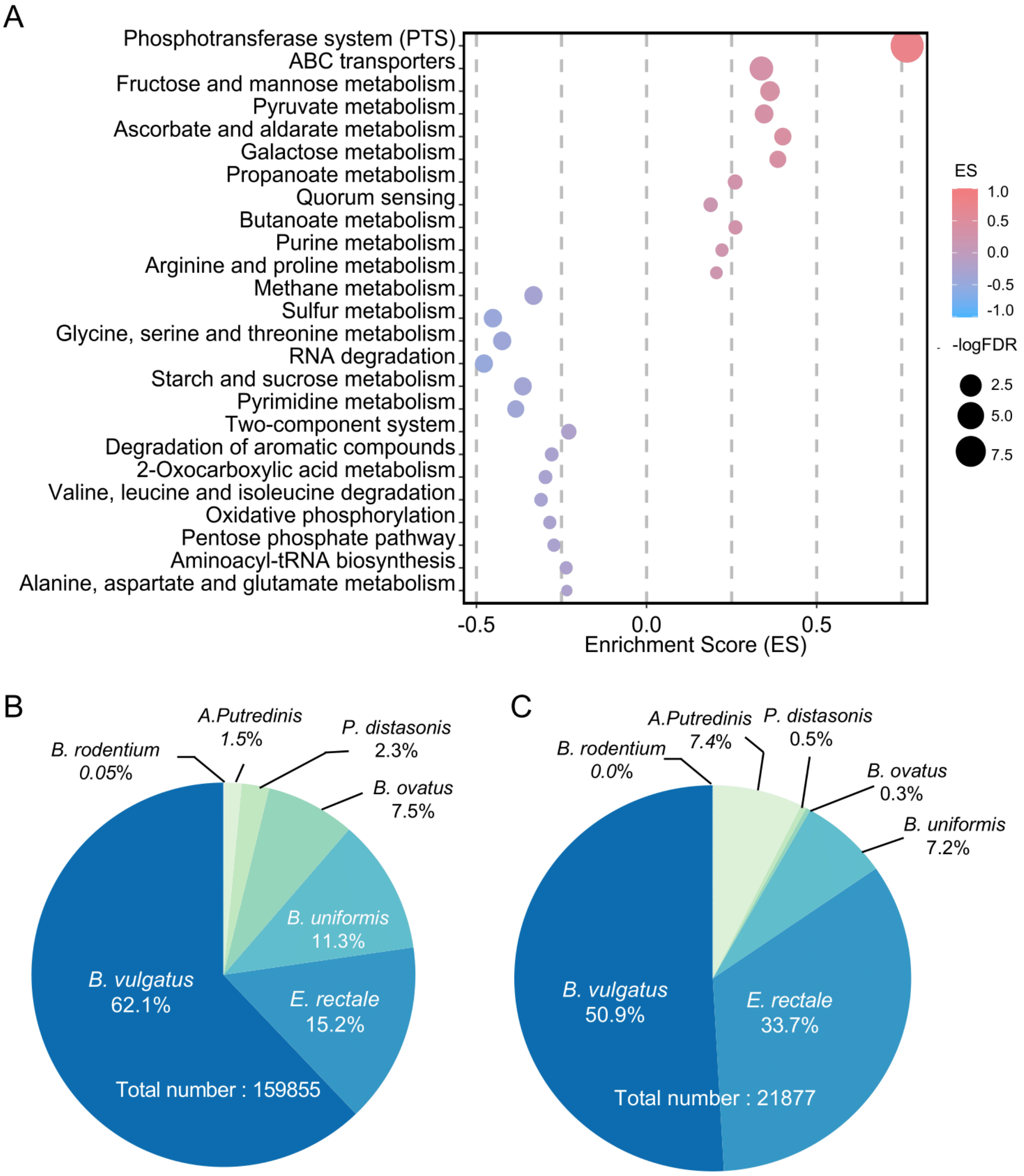


**Fig. S3. The gene set enrichment analysis of microbial genes, and distributions of annotated SNVs in species.**

**A** Dot plot of GSEA. The dot size and color represent the significance and the enrichment score of the enriched pathways, respectively. The ES>0 represents the up-regulation of pathway, and vice versa. **B-C** Pie chart showing the percentage of all annotated SNVs (B) and differential SNVs (C) in each species.

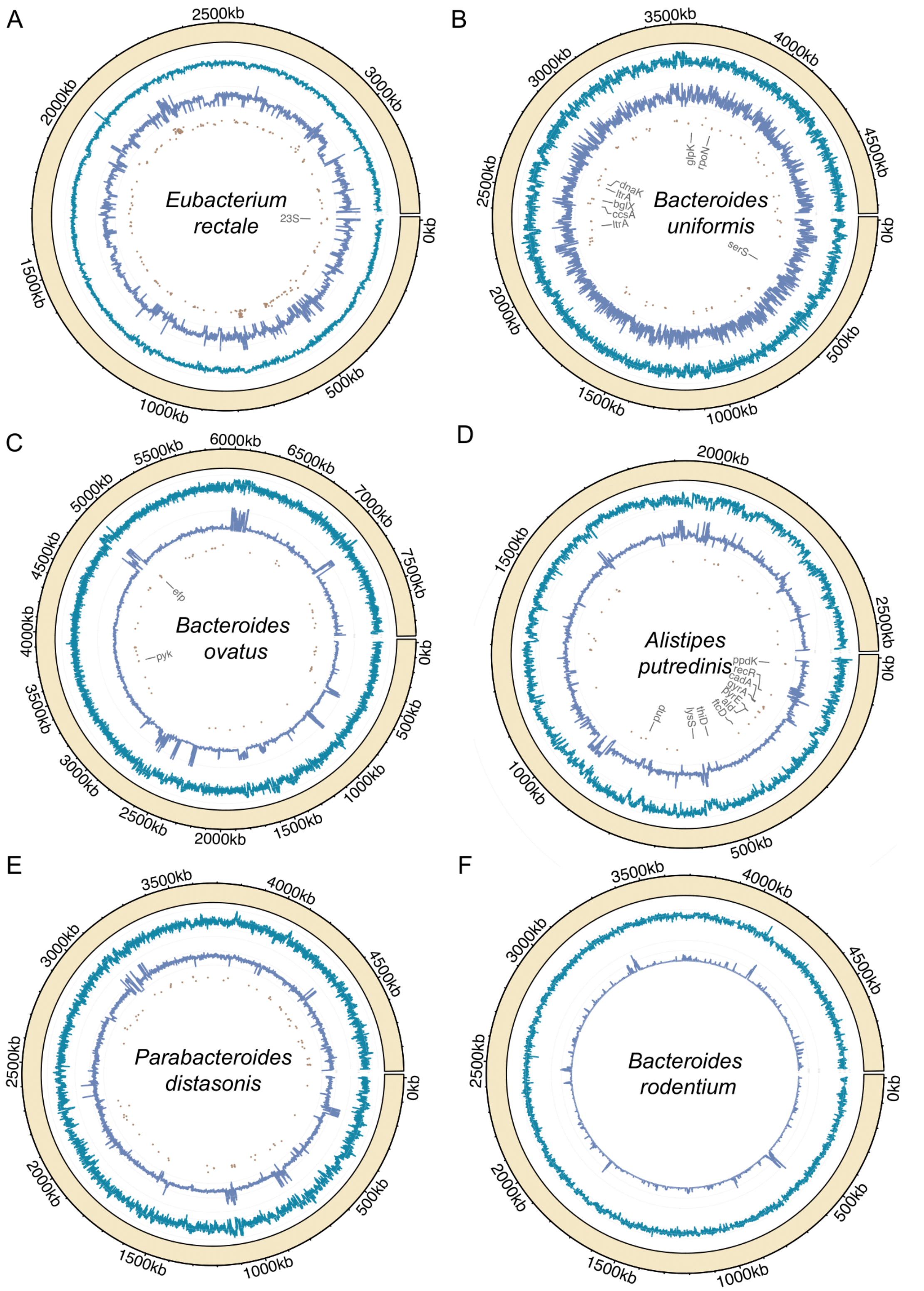


**Fig. S4. The annotated SNVs in the remaining six species with sufficient coverage. A-F** Chord diagram shows the distributions of annotated SNVs in *Eubacterium rectale* (A), *Bacteroides uniformis* (B), *Bacteroides ovatus* (C), *Alistipes putredinis* (D), *Parabacteroides distasonis* (E) and *Bacteroides rodentium* (F) genomes, respectively.

The outer circle represents the genome of species; the inner circles represent the GC-content (dark-green lines), sequencing depth (purple lines) and sites of differential SNVs (brown points) in the genome, respectively.

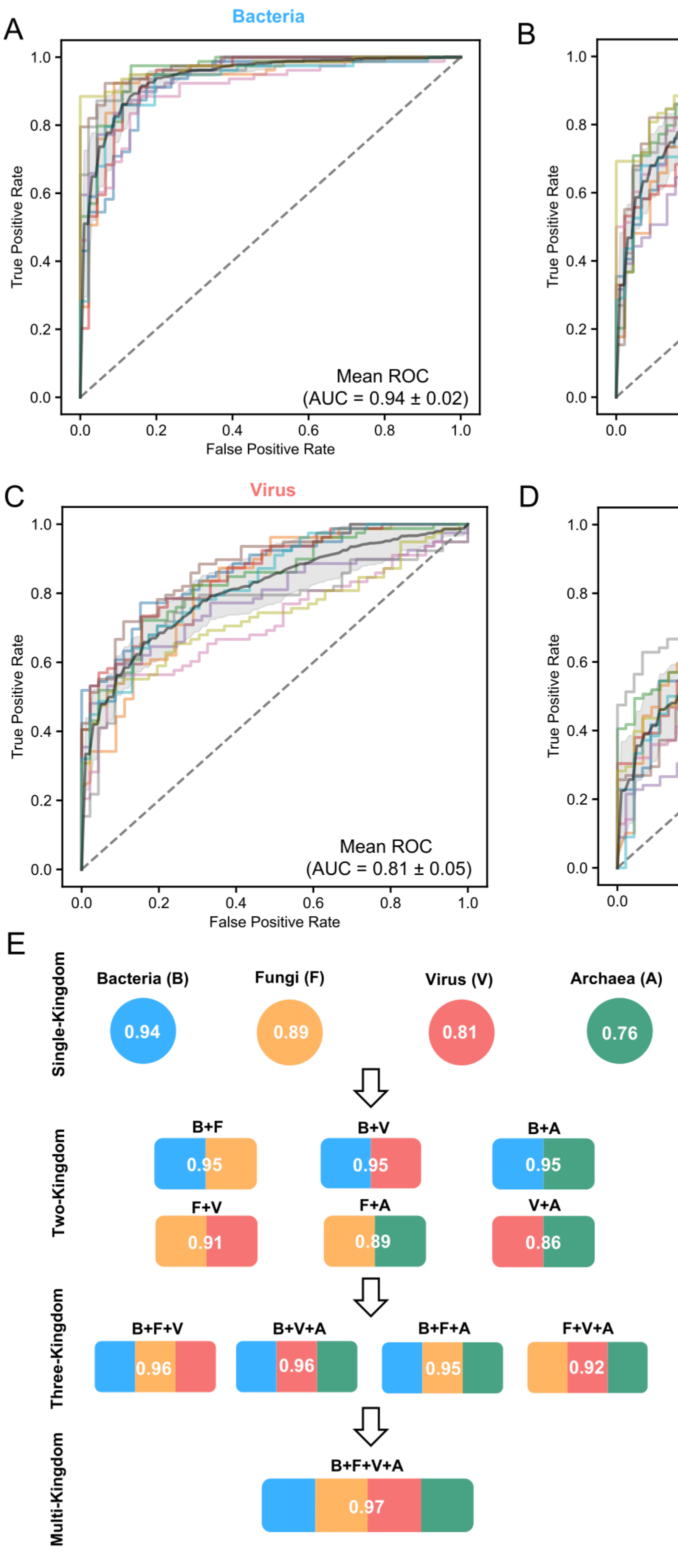

**Fig. S5. The diagnostic capabilities of multi-kingdom species models evaluated by cross-validation. A-D** ROC curves from ten-fold cross-validations of species model with bacterial (A), fungal (B), viral (C) and archaeal (D) signatures. **E** Average AUCs in cross-validation of models with single-, two-, three- and four-kingdom signatures.

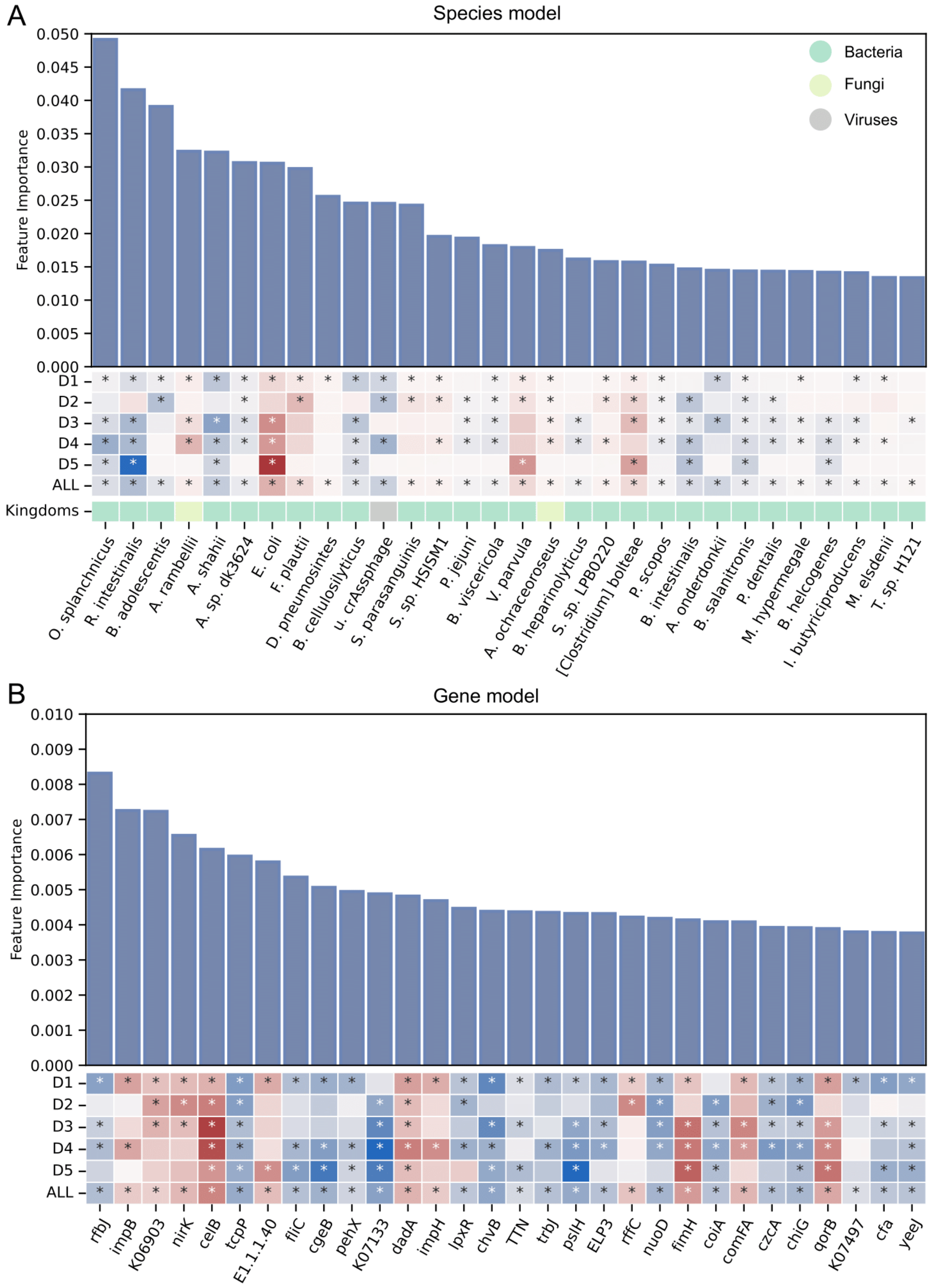


**Fig. S6. The top-ranking feature importance of species- and gene-models. A-B** Bar plot shows the feature importance values of species- (A) and gene-models (B); heatmap shows the increased (red) or decreased (blue)

abundances of the features in individual cohorts; the background color of ‘kingdoms’ bar represents the kingdoms of species signatures; * indicates statistical significance.

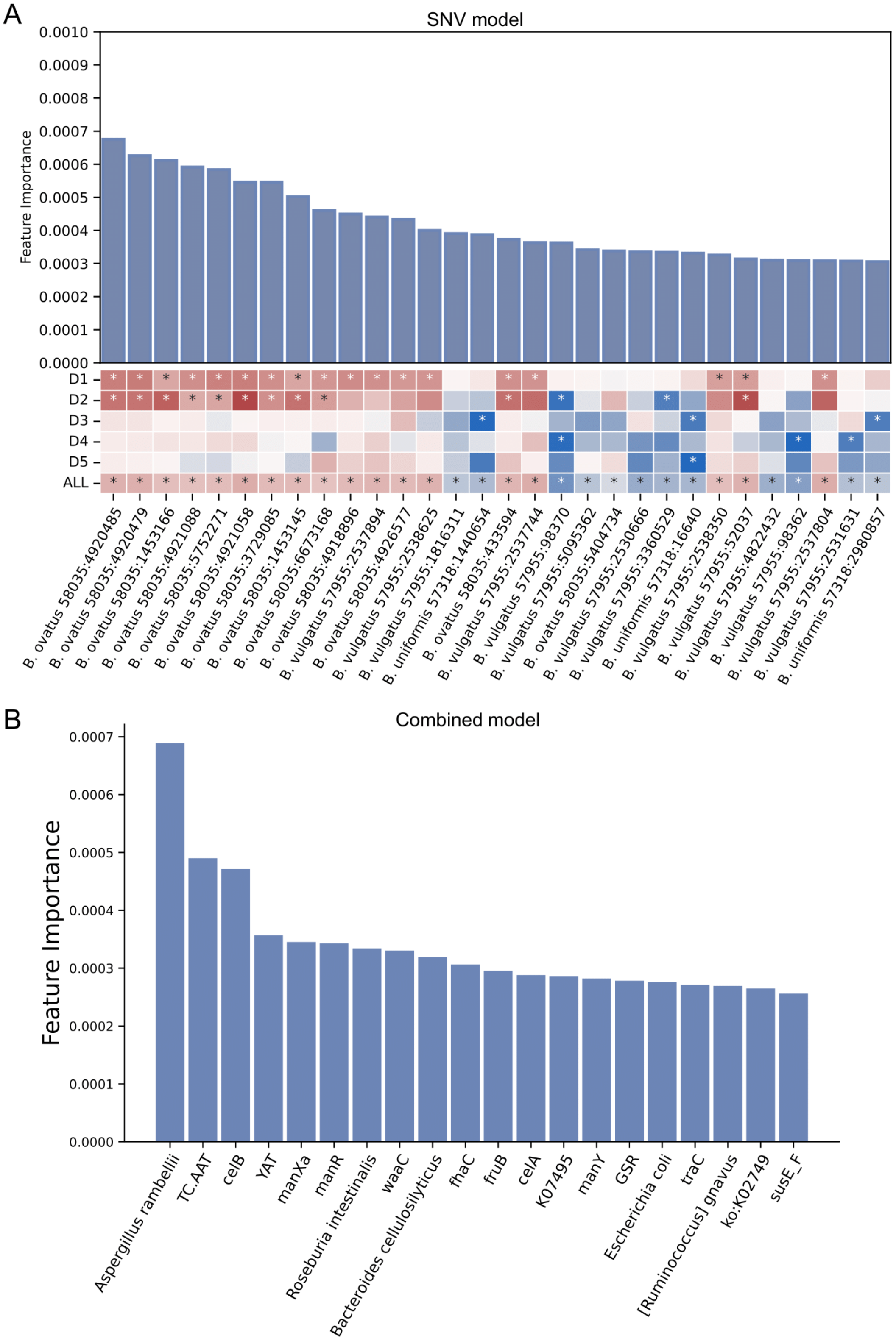


**Fig. S7. The top-ranking feature importance of SNV- and combined models. A** Bar plot shows the feature importance values of SNV-model; heatmap shows the increased (red) or decreased (blue) abundances of the features in individual cohorts; * indicates statistical significance. **B** Bar plot shows the feature importance values of combined model with multidimensional signatures.

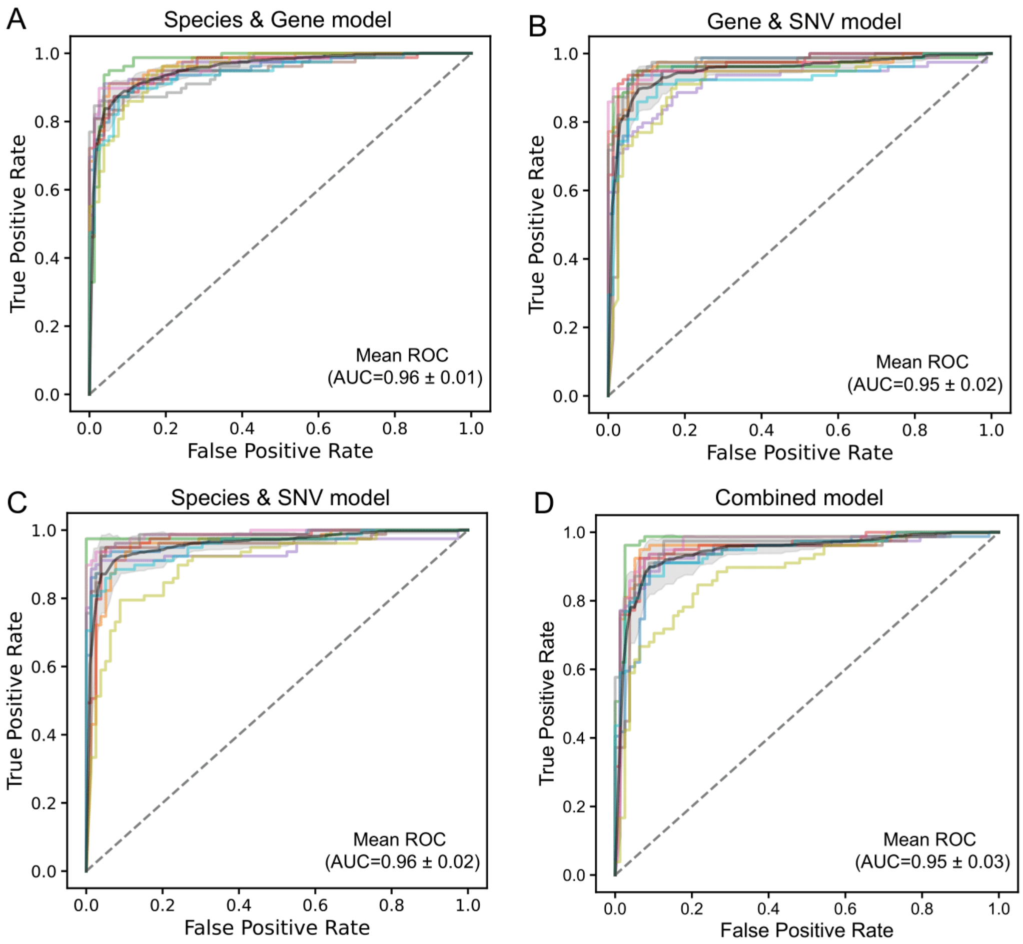


**Fig. S8. The diagnostic capabilities of multidimensional models evaluated by cross-validation. A-D** ROC curves in ten-fold cross-validation of models with species and gene- (A), gene and SNV- (B), species and SNV- (C), and combined multidimensional (D) signatures.

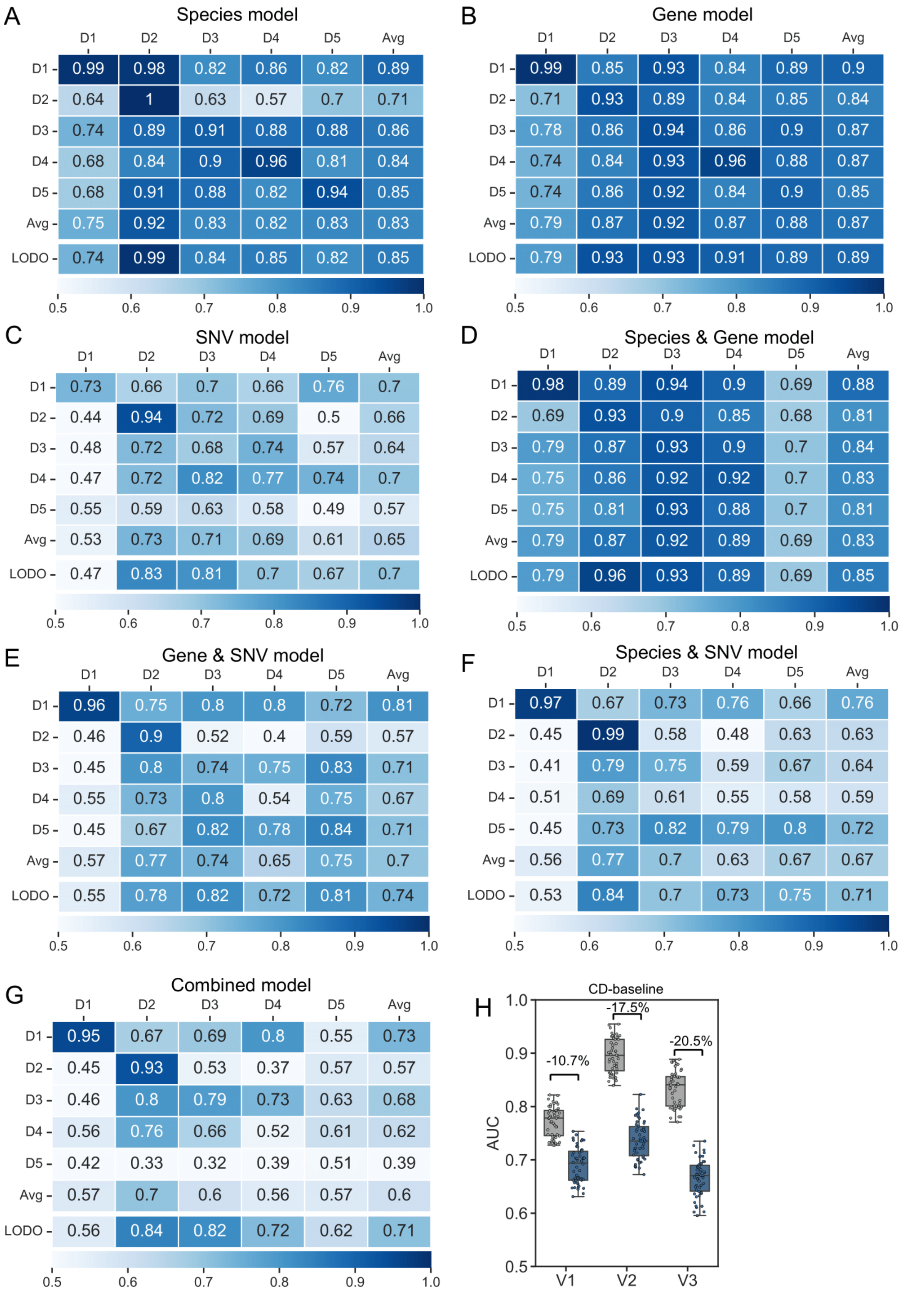


**Fig. S9. Internal validations of multidimensional models and AUC baseline of specificity evaluation. A-G** AUCs of internal validations of species- (A), gene- (B), SNV- (C), species and gene- (D), gene and SNV- (E), species and SNV- (F), and combined multidimensional (G) models, are listed. Each number in rows D1-D5 is the resulting AUCs of validation with the cohort specified by the row-tag as the training cohort, while all other cohorts combined as validation cohort; numbers in row Avg are mean values of the D1-D5; numbers in row LOCO are the resulting AUCs of validation with the cohort specified by the column tag as the validation cohort, while all other cohorts combined as the training cohort. **H** Plotted are the baseline values of the altered AUCs when adding CD samples in the three independent validation cohorts (V1, V2, V3). Gray and colored bars are the AUCs after adding control and case samples from the CD disease cohort, respectively.

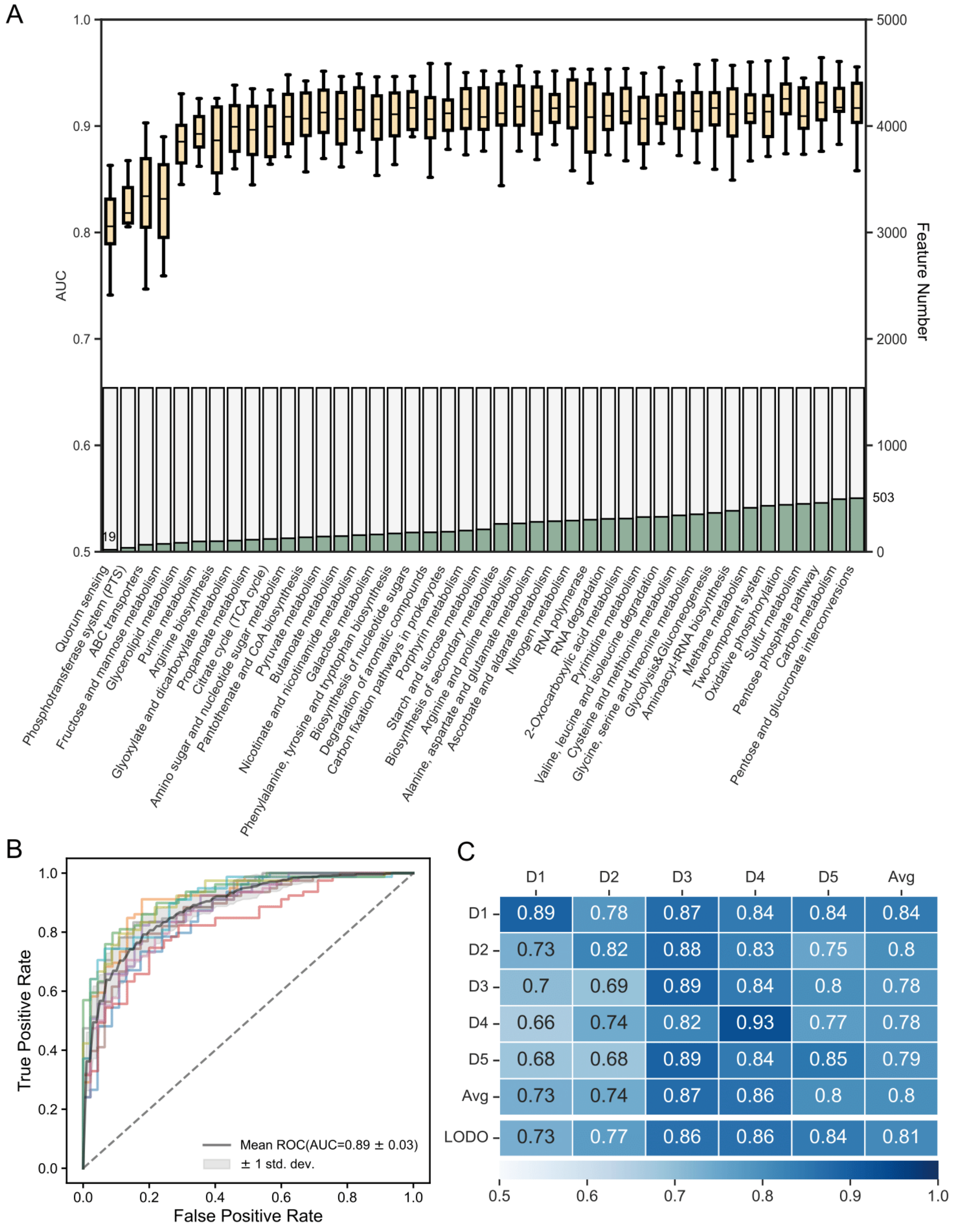


**Fig. S10. The recursive pathway elimination and diagnostic performances of sub-models. A** The box plot shows the AUCs of models in recursive pathway elimination analyses with the validation dataset. The bar plot shows the numbers of used features in each sub-model and total features in original gene models. **B** The ROC curves in ten-fold cross-validation of the sub-models. **C** The internal validations of the sub-models constructed with gene sets of quorum sensing, PTS, ABC transporters, fructose and mannose metabolism and glycerolipid metabolism. Each number in rows D1-D5 is the resulting AUCs of validation with the cohort specified by the row-tag as the training cohort, while all other cohorts combined as validation cohort; numbers in row Avg are mean values of the D1-D5; numbers in row LOCO are the resulting AUCs of validation with the cohort specified by the column tag as the validation cohort, while all other cohorts combined as the training cohort.

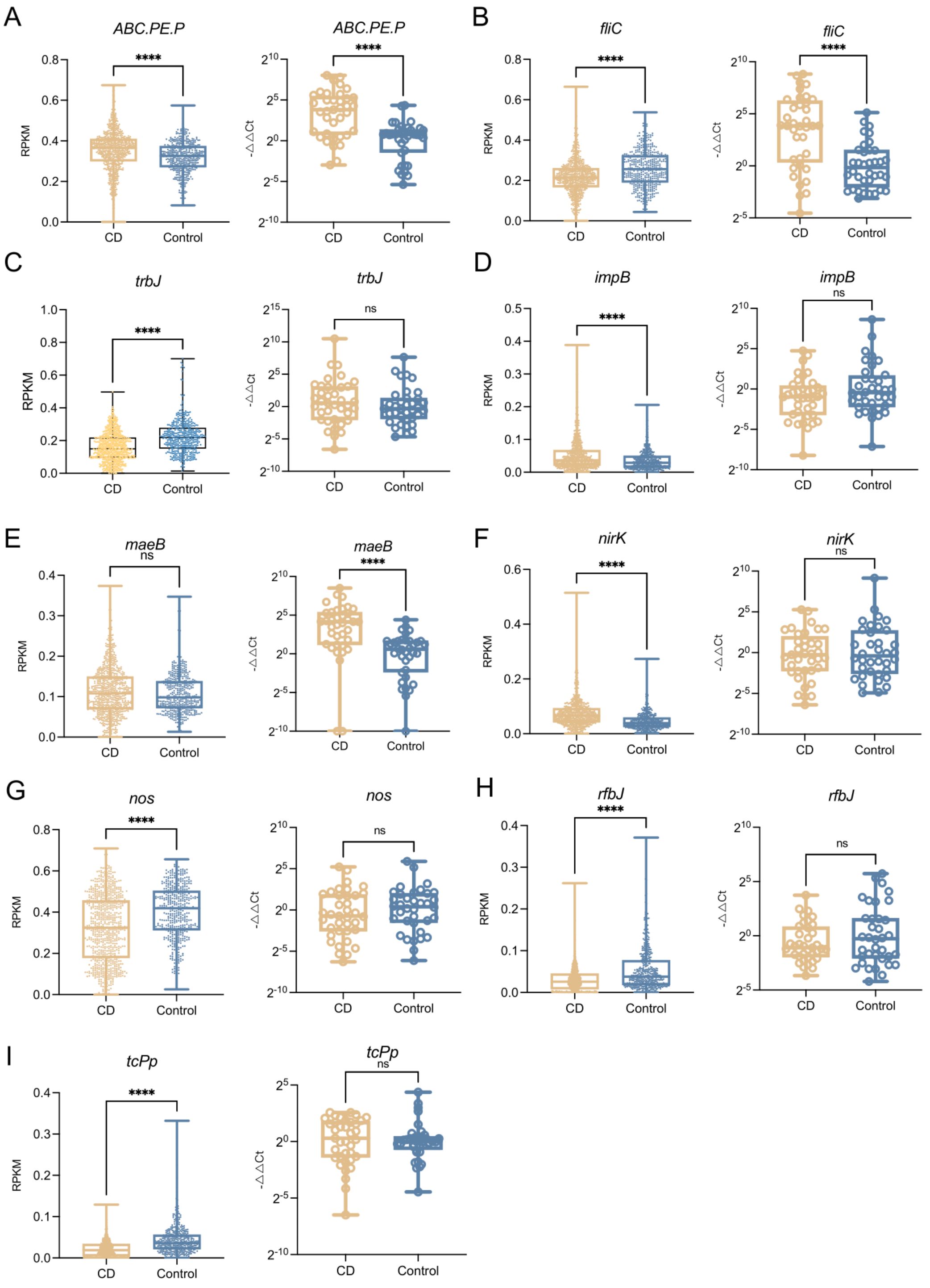


**Fig. S11. The abundance of microbial genes in qRT-PCR analysis. A-I** The abundance of *ABC.PE.E* (A), *fliC* (B), *trbJ* (C), *impB* (D), *maeB* (E), *nirK* (F), *nos* (G), *rfbJ* (H) and toxin coregulated pilus biosynthesis protein P (*tcPp)* (I) in the gut microbiome of CD and controls according to metagenomic data (left) and qRT-PCR (right; N =

37, CD; N = 36, control), respectively. Data are presented as mean ± standard deviation. *$P < 0.05$, **$P < 0.01$, ***$P < 0.001$ and ****$P < 0.0001$.

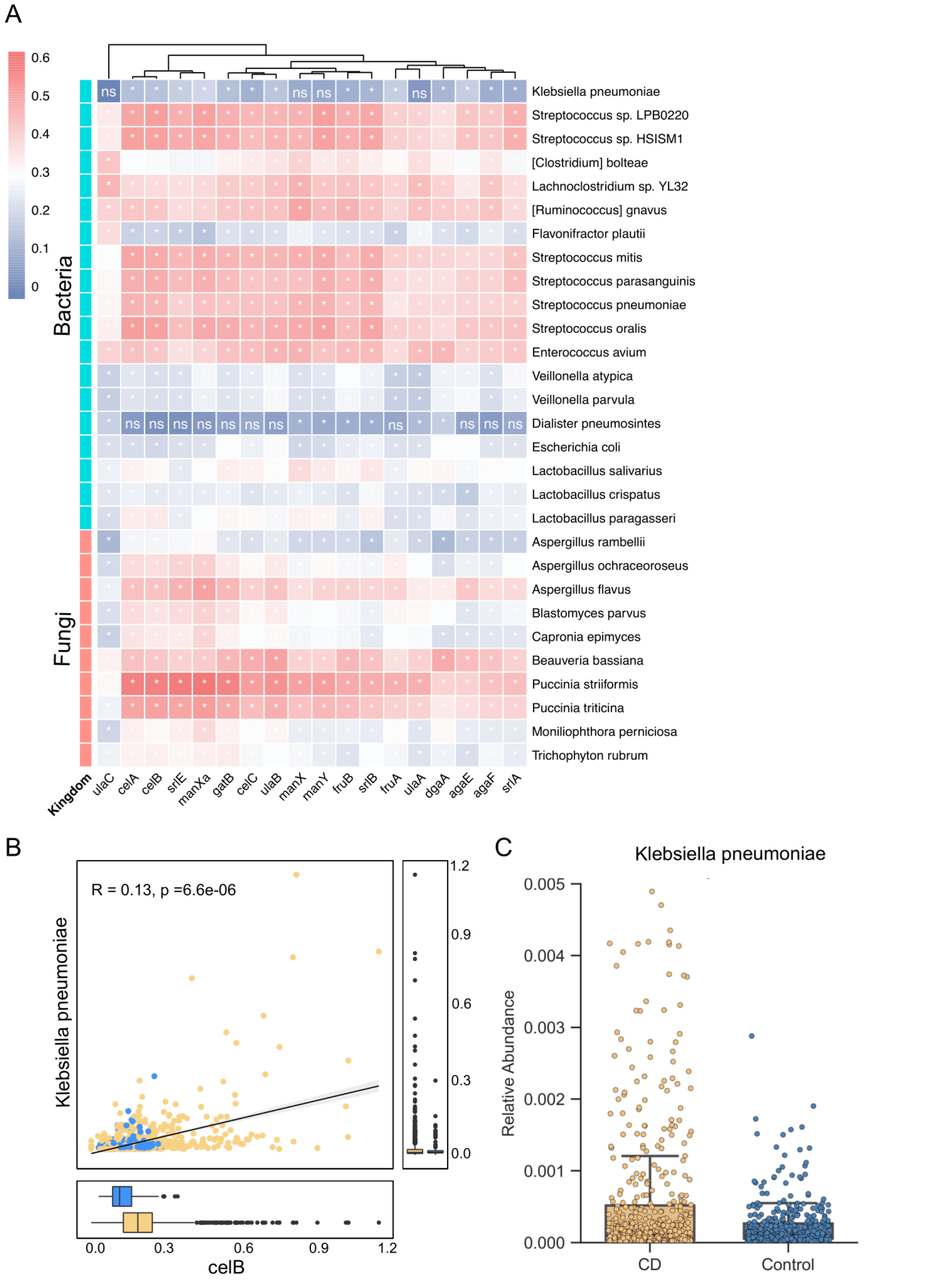


**Fig. S12. The cross talk between microbial genes and species. A** The correlations between genes in phosphotransferase system (PTS) and microbial species (bacterial species, green rows; and fungal species, orange rows). The color bar represents the ranges of correlation coefficients. The correlation significances are marked with asterisk if $P < 0.05$. **B** The linear regression between *celB and K. pneumoniae* in metagenomic data (Correlation = 0.13; $P = 6.6e\text{-}6$). **C** The relative abundance of *K. pneumoniae* in CD and controls.
